# Epidemiology of 376,383 arteriovenous fistula creations for hemodialysis over 9 years within the Brazilian Unified Public Health System and the Private Healthcare Sector

**DOI:** 10.1101/2025.09.02.25334945

**Authors:** Júlia Freire Castanheira de Paiva Ferreira, Thaís de Jesus Teani, Clara Sanches Bueno, Bruno Jeronimo Ponte, Felipe Soares Oliveira Portela, Marcelo Fiorelli Alexandrino da Silva, Marcelo Passos Teivelis, Miguel Cendoroglo Neto, Alexandre Fioranelli, Nelson Wolosker

**Affiliations:** Faculdade Israelita de Ciências da Saúde Albert Einstein - São Paulo, SP, Brazil; Hospital Israelita Albert Einstein - São Paulo, SP, Brazil; São Paulo University Medical School - São Paulo, SP, Brazil; Faculdade de Ciências Médicas da Santa Casa de São Paulo - São Paulo, SP, Brazil

**Keywords:** arteriovenous fistula, hemodialysis, public sector, private sector

## Abstract

**Background:** Chronic kidney disease (CKD) represents a growing public health challenge worldwide, particularly in low- and middle-income countries. In Brazil, where most patients with end-stage renal disease (ESRD) depend on the public healthcare system for dialysis and transplantation, ensuring adequate vascular access - among the options arteriovenous fistula (AVF) - is critical to sustaining treatment. Despite its clinical relevance, comprehensive national data on vascular access patterns across both public and private sectors have been historically limited.

**Objective:** This study aimed to examine trends in AVF confection for hemodialysis across Brazil from 2015 to 2023, assessing differences in frequency, geographic distribution and sectorial disparities between the public and the private healthcare sector.

**Methods:** A retrospective population-based analysis was conducted using anonymized data from national administrative databases: DATASUS (public sector) and D-TISS (private sector). The majority of AVF confection procedures were included. Statistical analyses considered regional adjustments and were performed using SPSS v.20, with significance set at *p* < 0.001.

**Results:** Over the nine-year period, 376.383 AVF procedures were recorded, with 90,57% occurring in the public sector. While the absolute number of AVF confectioners increased, the ratio of AVFs per 1,000 dialysis patients showed a declining trend. Regional disparities were evident, with the Southeast and South regions presenting higher procedure rates compared to other areas. The private sector consistently reported lower confection rates.

**Conclusion:** Although, separately, dialysis and AVFs rates have risen steadily in Brazil, the relative rate between than has decreasing trend over the years. The predominance of AVF procedures in the public system maintains higher when compared with the private sector.

## Introduction

Chronic Kidney Disease (CKD) is a significant global health concern, affecting approximately 10% to 13,4% of people worldwide. (1) (2) CKD is defined by structural or functional abnormalities in the kidneys that last for more than three months. If left untreated, CKD can progress to end-stage renal disease (ESRD). At this point, patients may require renal replacement therapies such as hemodialysis (HD) or peritoneal dialysis and may also be eligible for kidney transplantation (KT). (3)

In 2010, approximately 4.9 to 9.7 million people required kidney replacement therapy (KRT). (4) This scenario is particularly common in low- and middle-income countries (LMIC), where many patients with terminal CKD do not receive KRT. (5) In contrast, some countries, such as the United States, reported some of the highest rates of replacement therapy in the world; however, these rates have declined since 2017. (4) (6)

In 2023, Brazil reported a total of 157,357 dialysis patients. Of these, 96.2% received HD, while 3.8% underwent peritoneal dialysis. (5) From 2015 to 2023, a total of 46.272 kidney transplants were performed in the country, averaging 5,141 cases per year. The number of transplants ranged from a minimum of 4,395 to a maximum of 5,834 cases per year. (7)

To enable effective hemodialysis and improve patient survival, reliable vascular access is essential. The two main options for chronic treatment are arteriovenous fistulas (AVFs) and tunneled cuffed central venous catheters (TC-CVADs). (8) The selection of vascular access must be tailored for each patient. However, U.S. guidelines suggest that approximately 66% of patients undergoing maintenance hemodialysis should utilize AVs. (9) In cases where urgent dialysis is necessary and an AVF cannot mature quickly enough, it is appropriate to use a tunnelled central catheter or a short-term catheter. Plans should be in place to remove these catheters once the fistula becomes functional.

TC-CVADs are commonly used for the immediate initiation of dialysis or while patients are awaiting transplantation. (10) Consequently, their usage has become increasingly prevalent, often exceeding what is clinically appropriate. These catheters provide temporary access for patients who will eventually benefit from an AVF, either while waiting for the procedure or during the maturation process of the fistula. However, they carry the same costs associated with complications as all types of catheters. (11) Although the conversion to fistulas has been gradually increasing, it still lags behind the use of catheters, especially among older patients in the US. (10)

In Brazil, with a population of approximately 200 million people (12), around 72% rely on the publicly funded Unified Health System (SUS). The remainder is mainly covered by employer-sponsored private insurance (about 23%), while a small minority (less than 5%) pays for healthcare services out of pocket. (13) A recent study conducted in 2024 among public healthcare users—representing nearly 140 million individuals—identified a gradual reduction in the use of AVFs, accompanied by a growing reliance on TC-CVADs. (14)

The public healthcare system in Brazil has established a digital platform that collects data on procedures performed in public hospitals across the country. Recently, the private healthcare sector has also begun making its data available to the public, adopting practices similar to those of the public sector. By aggregating data from both sectors, it has become possible to conduct a comprehensive nationwide assessment of health in Brazil. This task was previously challenging due to limited data availability, given the country’s vast size.

This study aimed to analyze national data from both the public and private healthcare systems in Brazil between 2015 and 2023, regarding AVFs for hemodialysis. The objectives were to assess AVF rates, associated costs, and differences across regions, age groups, and sexes within both the public and private healthcare sectors. To our knowledge, this is the first study to describe and analyze such an extensive dataset in a country of continental dimensions. The findings provide comparative evidence that can inform the development of management strategies for CVD tailored to the Brazilian context.

## Materials and Methods

This is a retrospective, cross-sectional and population-based study, involving 376,383 procedures of patients with ESRD who underwent the creation of AVF across Brazil from 2015 to 2023. It offers a comprehensive national perspective on vascular access for hemodialysis over nine years.

The research protocol was reviewed and approved by the institutional ethics committee under protocol number 4324-20. Since all data were obtained from fully anonymized databases, the requirement for informed consent was waived, ensuring compliance with ethical standards while maintaining patient confidentiality.

The data were extracted from the TabNet platform of the Department of Informatics of the Unified Health System (DATASUS) and the Private Sector (D-TISS). (15-17) DATASUS is a digital platform that collects data from procedures performed in public hospitals across Brazil. The notification in this database is required for the procedure to be eligible for payment. The D-TISS is managed by a private system that provides access to procedure data. Overall population samples were derived from the Brazilian Institute of Geography and Statistics (12), the Brazilian Dialysis Census (5) (18-22) as well as DATASUS and D-TISS. (15–17)

This study analyzed procedures related to AVFs – confection - within the public and private systems, encompassing most of the hemodialysis services across Brazil over 9 years. The extracted data included the prevalence of sex, age, number of procedures performed in-hospital and ambulatory, and geographic region. The amount of dialysis per region was extracted from the Census (an estimated rate of HD over a sample of each region) and stratified by the region’s population. Regarding the overall analysis of the country, it was able to extract from the Census the total number of dialysis patients and the percentage that represents hemodialysis. In addition, with the percentage of HD financed by the public and the private systems, it was able to estimate the hemodialysis in each sector.

Procedure codes employed during selection and extraction on DATASUS to AVF confection are 0406020086 (for access), 0418010021 (with autologous graft), 0418010030 (for hemodialysis) and 0418010013 (with polytetrafluoroethylene (PTFE) graft). As for D-TISS AVF confection 30913144 (for hemodialysis) and 30908027 (with graft).

Automated data retrieval was performed using Python (version 2.7.13; Beaverton, OR, USA) within a Windows 10 environment. Specific fields were identified, and tables were modified using Selenium WebDriver (version 3.1.8; SeleniumHQ) in conjunction with the Pandas library (version 2.7.13; Lambda Foundry, Inc., PyData). All information extracted was organized and compiled on Microsoft Office Excel 2016® (v. 16.0.4456.1003, Redmond, WA, USA).

We initially analyzed the estimated number of hemodialysis patients in both public and private services between 2015 and 2023. We also examined the estimated creation of AVF in these services and the rate of AVF creation per thousand dialysis patients. In addition, the difference between age and sex, broken down by region, and the overall AVF creation, also categorized by region

### Statistical analysis

The statistical evaluation was conducted with SPSS version 20.0 (IBM Corp, Armonk, NY). The evaluation of the relationship between procedure rates and region was performed using generalized linear models with a Gamma distribution. For sex and age, generalized models with a Poisson distribution were used, incorporating the number of procedures as an offset. A p-value of less than 0.001 was regarded as statistically significant.

## Results

During the nine-year study period, a total of 376,383 AVF procedures were performed, along with 1,144,984 HD procedures. The creation of AVFs is more prevalent among males, revealing a significant difference in gender distribution between the public and private healthcare sectors (p<0.001). In the public system, the rates are approximately ten times higher for both sexes compared to the private sector.

Additionally, there is statistical evidence indicating variations in fistula distribution by age group across these healthcare systems, as shown in Table 1. Younger individuals (aged 20-69 years) are more commonly found in the public healthcare system, while older age groups (70– 79 and over 80 years) are more frequently seen in private facilities.

**Table 1:** AVF confection according to the age on Public and Private Systems.

| Age | Public |  | Private |  | Total |  |
| --- | --- | --- | --- | --- | --- | --- |
|  | N | (%) | N | (%) | N | (%) |
| <14 | 725 | 0,21% | 117 | 0,33% | 842 | 0,22% |
| 15-19 | 2881 | 0,85% | 194 | 0,55% | 3075 | 0,82% |
| 20-29 | 16551 | 4,85% | 1155 | 3,26% | 17706 | 4,70% |
| 30-39 | 32216 | 9,45% | 3149 | 8,88% | 35365 | 9,40% |
| 40-49 | 54089 | 15,87% | 4369 | 12,32% | 58458 | 15,53% |
| 50-59 | 82584 | 24,22% | 6688 | 18,86% | 89272 | 23,72% |
| 60-69 | 88471 | 25,95% | 8341 | 23,52% | 96812 | 25,72% |
| 70-79 | 49760 | 14,60% | 7381 | 20,81% | 57141 | 15,18% |
| >80 | 13638 | 4,00% | 2968 | 8,37% | 16606 | 4,41% |
| Not Informed | 0 | 0,00% | 1106 | 3,12% | 1106 | 0,29% |
| **Total** | **340915** | **100%** | **35468** | **100%** | **376.383** | **100%** |

Most HD procedures were conducted within the public healthcare system, accounting for 80.68% of the total, as shown in Table 2. There was an increase in the demand for both public and private healthcare services. Notably, the HD rate per million inhabitants rose significantly, particularly in the private sector, which surpassed the public system in 2023.

**Table 2:** Estimated hemodialysis (HD) patients in Public and Private Services between 2015 to 2023.(5) (12) (18-22)

| Year | Public |  |  | Private |  |  |
| --- | --- | --- | --- | --- | --- | --- |
|  | D Patients | Population | Rate per million | HD Patients | Population | Rate per million |
| 2015 | 84.945 | 154.358.599 | 550,31 | 16.785 | 48.045.043 | 349,37 |
| 2016 | 93.789 | 157.552.513 | 595,29 | 19.210 | 46.319.412 | 414,73 |
| 2017 | 96.635 | 159.814.818 | 604,67 | 21.213 | 45.396.739 | 467,27 |
| 2018 | 98.550 | 161.386.087 | 610,64 | 24.637 | 45.142.951 | 545,76 |
| 2019 | 102.852 | 162.907.549 | 631,35 | 27.340 | 44.992.550 | 607,66 |
| 2020 | 109.397 | 164.004.412 | 667,04 | 24.668 | 45.160.477 | 546,23 |
| 2021 | 114.172 | 163.553.169 | 698,07 | 25.403 | 46.550.473 | 545,70 |
| 2022 | 117.720 | 162.944.585 | 722,45 | 28.880 | 47.918.398 | 602,70 |
| 2023 | 105.756 | 162.943.737 | 649,04 | 33.032 | 48.751.421 | 677,55 |
| Total | 923.816 | 1.449.465.469 | 637,35 | 221.168 | 418.277.464 | 528,76 |

The estimated number of AVF procedures performed in Brazil from 2015 to 2023, across both public and private healthcare services, is presented in Table 3. Our observations indicate that the public sector had AVF proportions nearly two to three times higher than those in the private sector. Adjusting rates (firstly, for hemodialysis over fistula) for better description, in the public sector, decreased from 40.6% in 2015 to a low of 33.5% in 2022, followed by a slight recovery in 2023. Conversely, in the private sector, the decline was more pronounced, dropping from 25.7% to 12.8% during the same period. Either way, the results continue to be more expressive and significant on public systems.

**Table 3:** Estimated AVF confection in Public and Private Services between 2015 and 2023.

| Year | Public |  |  | Private |  |  |
| --- | --- | --- | --- | --- | --- | --- |
|  | AVF | HD Patients | AVF over HD (%) | AVF | HD Patients | AVF over HD (%) |
| 2015 | 34518 | 84.945 | 40,64% | 4.310 | 16.785 | 25,68% |
| 2016 | 34567 | 93.789 | 36,86% | 3.870 | 19.210 | 20,15% |
| 2017 | 35976 | 96.635 | 37,23% | 4.584 | 21.213 | 21,61% |
| 2018 | 37934 | 98.550 | 38,49% | 3.686 | 24.637 | 14,96% |
| 2019 | 40366 | 102.852 | 39,25% | 3.807 | 27.340 | 13,92% |
| 2020 | 38489 | 109.397 | 35,18% | 3.161 | 24.668 | 12,81% |
| 2021 | 38389 | 114.172 | 33,62% | 3.820 | 25.403 | 15,04% |
| 2022 | 39500 | 117.720 | 33,55% | 3.996 | 28.880 | 13,84% |
| 2023 | 41176 | 105.756 | 38,93% | 4.234 | 33.032 | 12,82% |
| Total | 340.915 | 923.816 | 36,90% | 35.468 | 221.168 | 16,04% |

The number of AFVs in the public sector was nearly ten times higher than in the private sector, with an average of 38,389 procedures (CI 2,661; 11,551) compared to 3,870 (CI 48; 202), respectively.

Table 4 presents the number of AVFs performed from 2015 to 2023, organized by region in Brazil and by type of service (public and private). In every region, the public sector consistently outperformed the private sector in terms of AVF procedures, with particularly significant differences in the North and Northeast regions.

**Table 4:** AVF confection divided by regions.

| Year | North |  | Northeast |  | Midwest |  | Southeast |  | South |  |
| --- | --- | --- | --- | --- | --- | --- | --- | --- | --- | --- |
|  | Public | Private | Public | Private | Public | Private | Public | Private | Public | Private |
| 2015 | 1.643 | 14 | 9.480 | 188 | 2.636 | 137 | 15.436 | 1.132 | 5.323 | 208 |
| 2016 | 1.575 | 22 | 9.714 | 251 | 2.564 | 175 | 15.326 | 1.507 | 5.388 | 216 |
| 2017 | 2.130 | 14 | 10.581 | 330 | 2.661 | 289 | 15.210 | 1.795 | 5.394 | 286 |
| 2018 | 2.034 | 24 | 10.950 | 306 | 2.910 | 206 | 16.442 | 1.790 | 5.598 | 297 |
| 2019 | 2.126 | 49 | 11.818 | 372 | 2.920 | 228 | 17.578 | 1.791 | 5.924 | 343 |
| 2020 | 2.063 | 35 | 10.687 | 233 | 2.914 | 211 | 17.010 | 1.584 | 5.815 | 270 |
| 2021 | 2.632 | 24 | 11.551 | 273 | 2.605 | 216 | 16.433 | 1.631 | 5.168 | 292 |
| 2022 | 2.599 | 24 | 11.219 | 350 | 2.878 | 208 | 17.199 | 1.801 | 5.605 | 285 |
| 2023 | 2.681 | 35 | 12.238 | 337 | 3.031 | 227 | 17.687 | 1.780 | 5.539 | 248 |
| **Total** | 19.483 | 241 | 98.238 | 2640 | 25.119 | 1897 | 148.321 | 14.811 | 49.754 | 2445 |

The adjusted model, which accounts for geographic region, demonstrated an annual increase in procedure rates, with an estimated rate growth ranging from 0,4 to 7,9% per year. Over the past nine years, the public healthcare system has shown substantial and significant improvements across all regions, illustrated in *Table 4*.

The rate of AVF creation per thousand dialysis patients in Brazil, stratified by region from 2015 to 2023, is shown in Table 5. Overall, there was either a reduction or fluctuation in rates across all regions over time, with an average annual decline of 1.6% (p = 0.01). Notably, the Northeast and North regions exhibited higher rates compared to the South and Southeast.

**Table 5:** Rate of AVF over per thousand patients in Dialysis divided by region.

| Year | North | Northeast | Midwest | Southeast | South |
| --- | --- | --- | --- | --- | --- |
| 2015 | 320,39 | 382,41 | 291,04 | 303,33 | 333,14 |
| 2016 | 262,17 | 361,74 | 283,46 | 278,46 | 310,02 |
| 2017 | 252,72 | 367,90 | 261,71 | 283,85 | 307,05 |
| 2018 | 252,65 | 374,16 | 298,94 | 281,66 | 318,53 |
| 2019 | 278,98 | 359,58 | 259,98 | 287,26 | 333,44 |
| 2020 | 397,02 | 317,75 | 240,90 | 263,75 | 293,79 |
| 2021 | 373,61 | 335,57 | 232,89 | 240,49 | 255,10 |
| 2022 | 268,27 | 309,51 | 269,23 | 228,88 | 272,20 |
| 2023 | 335,74 | 304,62 | 242,41 | 232,52 | 287,71 |

## Discussion

This retrospective, cross-sectional study examined the majority of Brazilian population. It analyzed a large sample of 376,383 procedures of ESRD patients who underwent AVF procedures in Brazil between 2015 and 2023, covering both the public and private sectors. It represents the largest and first nationwide cohort of AVF creation for hemodialysis, with robust epidemiological relevance for assessing vascular access across the entire country. The use of national databases—DATASUS for the public sector and D-TISS for the private sector—was effective in capturing nearly all procedures (excluding those who pay out-of-pocket), as reporting is mandatory for billing purposes. Integration of population data from the Brazilian Institute of Geography and Statistics (IBGE) and the Brazilian Dialysis Census enhanced the analysis, enabling demographic stratification and calculation of standardized rates.

Over the nine-year period, 1,144,984 HD procedures and 376,383 AVF procedures were performed in Brazil, mostly in the public sector (80.68% and 90.57% respectively), reaffirming its central role in ESRD care. While HD patients’ growth remained steady until 2022, a modest decline in the public sector occurred in 2023 (5), possibly linked to COVID-19 related mortality, (23) whereas the private sector continued to rise, surpassing the public in patients per million of dialyses.

The number of dialysis procedures has increased from 2022 to 2023, accounting for over 92.97% of the total HD volume. Over the past nine years, the number of hemodialysis is approximately 140,000 procedures. (5) Internationally, the United States recorded 639,883 new ESRD cases from 2015 to 2020, with a 3% decline in the use of AVF. (24) In Brazil, the incidence of dialysis rose from 45,852 in 2019 to 51.153 in 2023. (5) (18) In contrast, the number of AVF rose until 2019, dropped in 2020 due to COVID-19 (25), then rose again through 2023, though the AVF/HD ratio decreased, reflecting the increase of other approaches.

The data reveal significant differences between the public and private sectors in the use of AVFs for HD. The proportion of fistula over hemodialysis, in the public sector, ranged from 33.5% to 40.6%, while in the private sector, they fell sharply from 25.7% to 12.8%. Despite adjustments for population size, public rates remained higher, though the gap narrowed. This disparity suggests that, while fistulas are widely recognized as the preferred treatment due to their ability to reduce the risks of infection, hospitalizations, and costs, the private sector is more dependent on catheters. This reliance may be attributed to factors such as patient clinical profiles, institutional preferences, or barriers to accessing vascular surgery (9).

Even after adjusting for population size, AVF rates remained higher in the public sector, though the gap has decreased. Despite the overall increase in AVF creation, the percentage of AVFs relative to the total number of HD patients has declined, particularly in the private sector, raising concerns about prolonged catheter use and its poorer outcomes. This trend is concerning from a clinical standpoint, as it suggests a longer duration of central venous catheter use, which is associated with poorer outcomes. (14) Similar trends occur in countries with ageing dialysis populations and more comorbidities (24, 26), though international guidelines (KDOQI, ERA-EDTA) continue to recommend establishing definitive access whenever possible. (5,14,18– 22,27–29) In contrast, Japan, Italy, and Germany report AVF rates above 60–70%, far higher than in Brazil, particularly in the private sector, underscoring room for improvement. Adapting initiatives like the Fistula First campaigns in the United States, tailored to fit the Brazilian context, could help increase the rate of access to definitive care.

During the study period, 376,383 AVFs were reported as newly established, though these numbers may not represent unique patients. A significant number of these entries did not include detailed classifications regarding the specific type of fistula created. Additionally, published evidence indicates substantial variability in AVF patency outcomes, which are often influenced by various clinical and demographic factors, such as patient age, comorbid conditions, and the anatomical type of fistula used. (30) In Brazil, two national studies have shown that over 50% of patients required the procedure more than once. (31) (32) Due to the anonymous nature of the dataset, this study could not assess individual frequency or complications.

Regional analysis revealed that AVFs in the public sector significantly exceeded those in the private sector, particularly in the North and Northeast, where private involvement is relatively low compared to other regions. The Southeast had a higher absolute volume of procedures but one of the lowest AVF rates per thousand dialysis patients, indicating greater reliance on long-term catheters. Overall, AVF rates displayed declines or fluctuations across regions, with the highest rates in the Northeast and North, and the lowest rates in the South and Southeast, likely reflecting differences in vascular surgery infrastructure, regional health policies, and patient demographics. (33) In 2020, a peak of 397 AVFs per thousand HD patients was observed in the North. This increase may have been driven by specific initiatives, such as surgical campaigns or service reorganizations. At the same time, several regions saw declines, likely due to the suspension of elective procedures during the pandemic. AVFs were more frequent in men, with a statistically significant difference observed in both sectors (p < 0.001). In terms of age, younger patients (20–69 years) were predominantly treated in the public sector, whereas the private sector had a higher proportion of elderly patients (over 70 years). This age distribution may partly explain the lower AVF rates in the private sector, as older or comorbid patients are often less suitable for definitive vascular access.

Public healthcare performs far more HD and AVF procedures than the private sector, likely reflecting a higher prevalence of ESRD among its users, linked to poorer health conditions or limited access to healthcare. Additionally, many private patients seek these services in public facilities, a trend that has already been reported. (33)

In summary, although the public sector plays a dominant role and there has been overall growth in AVF creation in Brazil, the proportion of definitive access among HD patients is declining, particularly within the private sector, exposing regional and sectoral disparities that underscore the need for targeted interventions to ensure equitable access to care.

Continuous monitoring and robust databases are crucial for evaluating the impact of policies and informing effective strategies. Advanced vascular access planning for CKD is crucial in increasing the proportion of patients who start and maintain HD with definitive access. The decline in the AVF/HD ratio emphasizes the importance of implementing measures such as early referral to vascular surgery, training multidisciplinary teams, establishing national monitoring of indicators, and adopting international benchmarks. Moreover, policies must also address local demands, such as increasing vascular surgeon availability and specialized centers in underserved areas.

### Limitations

The study’s cross-sectional design does not permit causal inferences, thereby limiting its findings to associations and temporal trends. The lack of individual clinical data — such as comorbidities, disease stage, duration of dialysis, and reasons for choosing dialysis—further restricts the ability to account for specific risk factors and clinical contexts.

Reliance on secondary administrative databases may introduce inconsistent entries, while anonymization prevented patient-level longitudinal tracking, preventing the evaluation of outcomes and comorbidity profiles. It is important to note that the data reflect the number of fistula procedures performed, rather than the number of patients who underwent these interventions.

In the private sector, reliance on a single procedure code increases the risk of overestimating the data and complicates the analysis. Moreover, individuals without private health insurance who pay out of pocket were not included in the private sector dataset, which may affect the generalizability of the results.

Another limitation is the absence of standardized procedural coding for catheter placement, particularly in the private healthcare sector, which prevented inclusion of catheter-related interventions. Consequently, an essential component of HD practice could not be assessed, restricting a more comprehensive evaluation of access modalities within the population.

## Conclusion

In Brazil, the number of dialysis and hemodialysis patients has risen over the nine-year study period. However, the ratio of AVFs to dialysis has decreased. A consistent pattern was observed: AVF procedures are more frequent in the public sector than in the private sector (per numbers of beneficiaries).

## Data availability statement

All data presented in this study can be made available upon reasonable request to the corresponding author.

## Authors’ contributions

Conception and design: BJP, JFCPF, NW

Analysis and interpretations: BJP, JFCPF, TJT, MFAS, MPT NW

Data collection: BJP, JFCPF, MFAS

Writing the article: BJP, JFCPF, TJT, MPT

Critical revision: BJP, MFAS, MPT, NW

Final approval of the article: NW

Statistical analysis: BJP, JFCPF, MFAS, TJT

Overall responsibility: BJP, JFCPF, MFAS, NW

